# MicroRNA signature predicts post-operative atrial fibrillation after coronary artery bypass grafting

**DOI:** 10.1101/2024.06.21.24309328

**Authors:** Srinivasulu Yerukala Sathipati, Tonia Carter, Deepa Soodi, Nwaedozie Somto, Sanjay K Shukla, John Petronovich, Glurich Ingrid, John Braxton, Param Sharma

**Author notes:** Corresponding author email addresses.

## Abstract

**Background:** Early detection of atrial fibrillation (AFib) is crucial for altering its natural progression and complication profile. Traditional demographic and lifestyle factors often fail as predictors of AFib, particularly in studies with small samples. This study investigated pre-operative, circulating microRNAs (miRNAs) as potential biomarkers for post-operative AFib (POAF) in patients undergoing coronary artery bypass grafting (CABG).

**Methods:** We used an array polymerase chain reaction method to detect pre-operative, circulating miRNAs in seven patients who subsequently developed POAF after CABG (cases) and eight patients who did not develop POAF after CABG (controls). The top 10 miRNAs from 84 candidates were selected and assessed for their performance in predicting POAF using machine learning models, including Random Forest, K-Nearest Neighbors (KNN), XGBoost, and Support Vector Machine (SVM).

**Results:** The Random Forest and XGBoost models showed superior predictive performance, with test sensitivities of 0.76 and 0.83, respectively. Differential expression analysis revealed four upregulated miRNAs—hsa-miR-96-5p, hsa-miR-184, hsa-miR-17-3p, and hsa-miR-200-3p—that overlapped with the AFib-miRNA signature. The AFib-miRNA signature was significantly associated with various cardiovascular diseases, including acute myocardial infarction, hypertrophic cardiomyopathy, and heart failure. Biological pathway analysis indicated these miRNAs target key signaling pathways involved in cardiovascular pathology, such as the MAPK, PI3K-Akt, and TGF-beta signaling pathways.

**Conclusion:** The identified miRNAs demonstrate significant potential as predictive biomarkers for AFib post-CABG, implicating critical cardiovascular pathways and highlighting their role in AFib development and progression. These findings suggest that miRNA signatures could enhance predictive accuracy for AFib, offering a novel, noninvasive approach to early detection and personalized management of this condition.

## Introduction

Post-operative atrial fibrillation (POAF) represents a prevalent complication following coronary surgery, bearing a twofold increase in cardiovascular mortality and morbidity compared to individuals maintaining normal sinus rhythm [1]. POAF manifests in paroxysmal, persistent, or permanent forms, with onset frequently occurring within the initial 5 days, peaking at 48 to 72 hours post-surgery [2]. Incidence rates, estimated to be between 10 to 50%, underscore its clinical significance [3, 4]. Importantly, POAF is linked to severe adverse outcomes, including an elevated risk of venous thromboembolism (VTE) and stroke due to hemodynamic instability, impaired ventricular filling, and compromised cardiac output. Moreover, POAF may contribute to long-term cardiac pathology, exacerbating heart failure and culminating in renal decline, increased mortality, and augmented healthcare costs associated with extended hospital stays and emergent adverse events [3].

In the context of coronary artery bypass grafting (CABG), POAF is not exclusive to the choice of on- or off-pump approaches and is hypothesized to result from reperfusion injury in susceptible individuals with predisposing risk profiles. Risk indices have been devised to stratify patients based on their individual risk profiles [3]. The intricate array of underlying factors contributing to POAF includes pre-existing cardiac and vascular pathologies, inflammatory and oxidative stress, genetic predisposition, and intra/post-operative factors triggering compensatory cardiac remodeling [2]. Vulnerable populations, such as those with congestive heart failure, mitral valve pathology, advanced age, or atrial interstitial fibrosis, are particularly prone to POAF development [5]. Patients with coronary heart disease, hypertension, and ventricular hypertrophy are at risk for permanent atrial fibrillation (AFib) [6].

Despite extensive research, the mechanisms leading to POAF remain inadequately understood. Animal models, *in vitro* studies, and clinical trials indicate oxidative injury, inflammatory processes, altered myofibrillar energetics, neurohormonal activation, and volume overload as potential contributing factors [7–9]. Additionally, autoantibodies targeting M2 muscarinic receptors, myosin, or heat shock proteins, and chronic infections like chlamydial and *Helicobacter pylori* have been proposed as contributors, although causality remains to be established [10, 11]. The relationship between AFib development independent of surgical exposure and the mechanisms involved in POAF remains unclear. Despite the development of risk indices based on clinical parameters, predictive biomarkers remain elusive. Current preventive measures, although diverse, have not substantially reduced POAF incidence following CABG. Recently, microRNAs (miRNA) have emerged as promising biomarkers, offering potential for predicting disease onset and outcomes[12, 13]. These stable, non-coding RNAs play a crucial role in gene expression regulation across various biochemical pathways [14]. A seminal study by Barth et al. in 2005 observed distinct gene expression patterns in patients with permanent AFib compared to those in sinus rhythm among patients undergoing open heart surgery for valve repair or CABG, suggesting the potential of miRNAs as indicators of POAF risk [15].

The stable expression of circulating miRNAs post-sample collection is a characteristic that suggests these molecules have the potential to serve as reliable biomarkers for predicting future disease events, e.g., POAF. Alterations in miRNA levels are intricately linked to the signaling cascades governing electrical remodeling and atrial fibrosis in the heart. Therefore, specific miRNAs and their quantitative profiles potentially could serve as indicative markers for a spectrum of clinical scenarios related to the heart, e.g., acting as either favorable or unfavorable indicators for the onset of AFib [16, 17]. Evidence indicates that miRNAs serve as predictive markers for AFib in patients with cardiovascular diseases. For instance, Galenko et al. (2019) identified circulating miRNAs capable of discerning individuals at a heightened risk of AFib, and observed an association between reduced expression of miRNA-21 and AFib [18]. Kiliszek et al. (2020) also illustrated significant differences in the levels of 34 miRNAs in sera from patients with AFib recurrence compared to those without AFib recurrence [19]. Additionally, Cao et al. (2021) identified seven upregulated and 13 downregulated differentially expressed miRNAs to distinguish patients with AFib from healthy individuals [20]. However, given the complexities associated with AFib, there remains a need to explore feasible biomarkers that can predict first occurrence of AFib to prevent adverse outcomes in patients with cardiovascular diseases.

This study aimed to investigate whether miRNAs can predict POAF following CABG. We used machine learning methods to identify a group of circulating miRNAs, measured pre-operatively, that could distinguish between patients who went on to develop POAF and those who maintained sinus rhythm following CABG. We also determined whether this group of miRNAs had known associations with cardiovascular diseases, tested for enrichment of these miRNAs in biological pathways, with an emphasis on enrichment in pathways related to cardiovascular disease, and identified the target genes of these miRNAs to form a miRNA-target gene interaction network. The results will provide insights into the target genes and biologically relevant pathways that are impacted during the emergence of POAF.

## Methods

### Study subject inclusion and exclusion criteria

The Internal Review Board (IRB) from Marshfield Clinic Health System approved this cohort study prior to data access and determined that the study posed minimal risk to participants.

Patients who were candidates for CABG at Marshfield Clinic Health System (MCHS) were identified by a MCHS cardiologist (JB) as he encountered them in his practice, by reviewing the scheduled surgeries of three MCHS cardiologists who performed CABG procedures, or by alerts from medical staff of emergent, unscheduled CABG procedures to be performed by the three MCHS cardiologists. Before the CABG procedure was performed, identified patients were screened for study eligibility by a review of electronic health record (EHR) data conducted by study staff. Inclusion criteria were age ≥ 18 years at time of screening, patient was an established MCHS patient, CABG surgery was to be performed at MCHS, and patient had no history of a prior CABG procedure. Exclusion criteria were patient had a previous history of atrial fibrillation, patient had a previous CABG performed, patient was in shock, patient had a previous heart valve surgery, patient had a myocardial infarction with balloon pump procedure within 72 hours prior to CABG, patient was scheduled for cardiac procedures in addition to CABG, patient received a blood transfusion within two weeks prior to CABG, patient had an infection requiring antibiotics, patient was unable to communicate effectively enough to report POAF symptoms following discharge, patient had a history of non-compliance, and patient or caregiver had personal or cultural beliefs regarding the patient’s condition or medical care which made it unlikely they would report symptoms of AFib after hospital discharge, potentially producing misclassification error among controls. A MCHS cardiologist (JB) performed a second review of the EHR data to verify that patients met study eligibility criteria.

### Participant recruitment

Following screening and before the CABG procedure, eligible patients were provided with a description of the study and asked to provide informed consent to participate in the study. A short interview was conducted with participants who provided informed consent to obtain information not readily available in the EHR, including history of blood transfusions, use of prescription and over-the-counter medications, previous surgeries, previous cardiac-related hospitalizations, family history of cardiac and other serious diseases, and consumption of caffeine, alcohol, and tobacco products. Study participants also provided a 10 mL blood sample prior to their CABG procedure.

### Participant follow-up and ascertainment of case-control status

Within 30 days following patient discharge after the CABG procedure, details of the CABG procedure were extracted from the EHR by MCHS cardiologists. The details included type of CABG surgery (on- or off-pump), time on pump, number of vena cannulas used, type of anesthesia, length of surgery, aortic clamp time, type and amount of blood products received (if any), lowest hematocrit, lowest body (bladder) temperature achieved, perioperative medications, diagnosis of AFib following CABG, and occurrence of infection or stroke while hospitalized.

Cases were defined as patients who developed POAF within 30 days after the CABG procedure. Controls were patients who remained in sinus rhythm for 60 days following the CABG procedure. A MCHS cardiologist contacted patients by telephone once 30 days had elapsed following the CABG procedure to ask the patients whether they had been diagnosed with AFib following the procedure. For patients who reported POAF, the cardiologist confirmed the diagnosis by reviewing EHR data or, if the patient was treated at another facility after CABG surgery, obtaining and reviewing medical records from other facilities. For patients without POAF, the telephone interview also determined if, within the 30-day period, patients had an acute cerebral vascular event, received blood products, had other cardiac procedures, felt fluttering or pain in the chest, or experienced dizziness. At 60 days after the CABG procedure, patients were interviewed by telephone again to inquire about their health status. The patients diagnosed with POAF within 30-days after CABG were asked whether they had an infection that required treatment with an antibiotic, an acute cerebral vascular event, had received any blood products, or had any cardiac procedures, and, if yes, whether these occurred before or after the AFib diagnosis. The patients who did not have AFib within 30-days after CABG were asked whether they had an acute cerebral vascular event, received blood products, had other cardiac procedures, felt fluttering or pain in the chest, or experienced dizziness in the 30-60 day period after the CABG procedure. Patient EHR data were reviewed by MCHS cardiologists to verify patient reports given in the telephone interviews, and the clinical data were used to categorize patients by case-control status.

### Processing of blood samples, extraction of miRNA, and measurement of miRNA expression

Blood samples were collected in 10 mL tubes that contained no anticoagulants. The blood was allowed to clot in the tubes for 1-2 hours at room temperature before centrifugation (3000 rpm for 10 minutes) at 4°C. miRNA was extracted from the serum collected after centrifugation using the miRNeasy Serum/Plasma Kit (Qiagen, Valencia, CA), according to the manufacturer’s instructions. The quality (A260:A280 ratio) of the extracted miRNA was measured using a NanoDrop™ spectrophotometer (ThermoFisher Scientific, Waltham, MA). The expression of a panel of 84 miRNAs was measured by reverse transcription quantitative polymerase chain reaction (qPCR) in the miRNA samples using the Human Serum & Plasma miScript miRNA PCR Array (Qiagen), following the manufacturer’s instructions, on a LightCycler 480 Instrument (Roche, Indianapolis, IN). The 84 miRNAs were selected by the manufacturer for inclusion on the panel because the serum expression levels of the miRNAs had been correlated with heart disease, liver disease, atherosclerosis, diabetes, and certain cancers in previous research. Delta cycling threshold (ΔCt) values for each miRNA were calculated by subtracting the average Ct value for six housekeeping small RNAs (*SNORD61*, *SNORD68*, *SNORD72*, *SNORD95*, *SNORD96A*, and *RNU6-2*) from the Ct value for the miRNA.

### Statistical analysis

Descriptive statistics are presented as medians and interquartile ranges (IQRs) for continuous variables, and as counts and percentages for categorical variables. Baseline characteristics were compared using the Wilcoxon test for continuous data and the Pearson χ2 test for categorical data. All statistical analyses were conducted using R version 4.3.1 (R Project for Statistical Computing) with the tidycmprsk and ggplot2 packages.

### Machine learning methods

We utilized standard machine learning algorithms to differentiate between AFib and control groups. The dataset was divided into 80% training (*n*=12) and 20% validation (*n*=3) subsets. A correlation-based feature selection method was applied to identify the top 10 miRNAs out of 84, with a correlation threshold set at 0.7. Random Forest, eXtreme Gradient Boosting (XGBoost), K-Nearest Neighbors (KNN), and Support Vector Machine (SVM) methods were employed for classification. The models were built using R libraries including ‘randomForest’, ‘xgboost’, ‘kNN’, and ‘caret’. The prediction performance of the models was evaluated using receiver operating characteristic (ROC) curves with the R package ‘pROC’.

The correlation coefficient between two miRNAs, *A* and *B* can be calculated using the following equation.

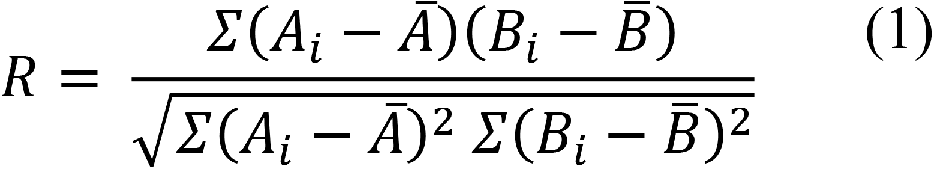

Where *Ai* and *Bi* are the individual data points for miRNAs *A* and *B* respectively.

*A̅* and *B̅* are the means of miRNAs *A* and *B* respectively.

### Differential expression analysis

In this study, we analyzed miRNA expression profiles from patients with AFib and control subjects using DESeq2. DESeq2’s statistical framework, based on a negative binomial distribution, accounts for variance-mean dependence and the biological variability typical of RNA-seq experiments. Although we have ΔCT values from PCR experiments, we opted to use DESeq2 for the statistical analysis. DESeq2 calculated normalized expression values, applied shrinkage estimators for variance stabilization, and conducted Wald tests to determine fold changes and associated p-values. To minimize false positives, a false discovery rate (FDR) adjusted *q*-value threshold of 0.05 was used.

### MiRNA-disease association and pathway analysis

We utilized miRNA-disease association information from HMDD v4.0 [21] and miRNet [22]. The AFib-miRNA signature and its disease associations were visualized using bar plots created with the ggplot2 (ver. 3.4.3) R package. The AFib-miRNA signature and its links to cardiovascular-specific diseases were displayed using an alluvial plot generated with the Alluvial R package. KEGG pathway analysis was conducted with DIANA-miRPath v4.0 [23]. For GO categories and Reactome pathways, we used both DIANA-miRPath v4.0 [23] and miRNet [22]. Specific pathways were visualized as bar plots using ggplot2 (ver. 3.4.3), and cardiovascular-specific pathways were displayed using chord diagrams created with the circlize R package.

### MiRNA-gene target interaction

To identify gene targets for each miRNA, we employed four different miRNA-gene target databases: miRWalk [24], miRNet [22], miRDB [25], and miRTarBase [26]. We ensured robust predictions by including only gene targets found in at least two of these databases. To further refine our results, we focused on gene targets supported by all four databases, ultimately identifying 50 genes associated with the AFib-miRNA signature. We used the ComplexUpset (version 1.3.3) R package to create upset plots.

## Results

### Patient characteristics

A total of 164 patients were screened for study eligibility. After applying the inclusion and exclusion criteria, 84 subjects were excluded due to various reasons, such as having a prior diagnosis of atrial fibrillation (*n* = 26), CABG (coronary artery bypass grafting) surgery performed previously or scheduled at another facility (*n* = 15 and *n* = 2, respectively), or other disqualifying factors as outlined in the flowchart (**Fig. 1**).

**Figure 1.**
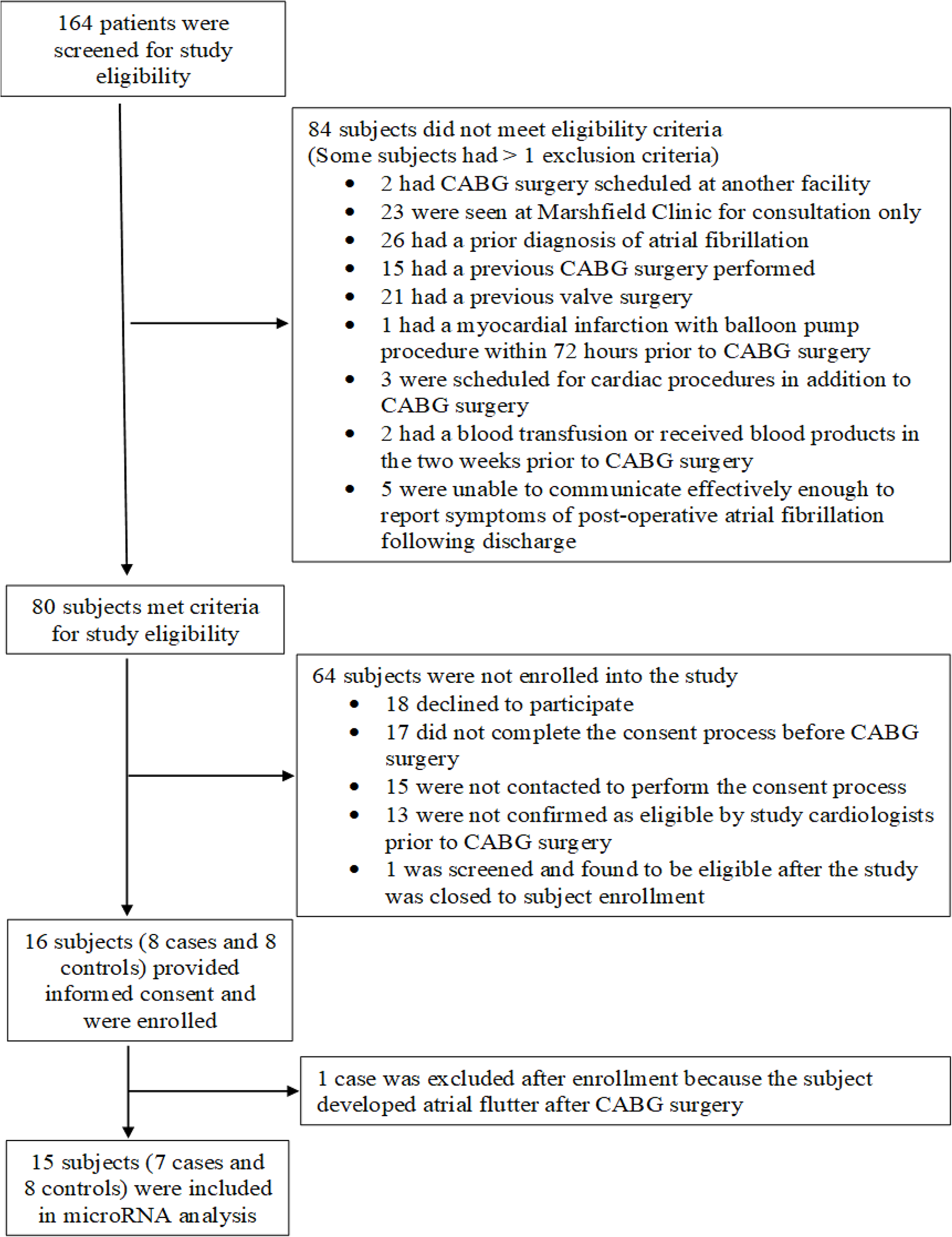
Flow chart of participant recruitment.

Of the remaining 80 subjects, 64 were further excluded due to declining to participate (*n* = 18), not completing the consent process before surgery (n = 17), or not being confirmed as eligible (*n* = 13). Ultimately, 16 subjects (8 cases and 8 controls) provided informed consent and were enrolled in the study. One case was excluded post-enrollment due to the development of atrial flutter following CABG surgery, resulting in a final cohort of 15 subjects (7 cases and 8 controls) for the miRNA analysis.

### Baseline Characteristics

The baseline characteristics of the study cohort are detailed in Table 1. The mean age at CABG surgery was 71.4 ± 7.7 years for cases and 68.4 ± 7.7 years for controls. Both groups consisted predominantly of male (cases: 85.7%, controls: 87.5%) and non-Hispanic white individuals (100% in both groups). The majority of participants in both groups were married (cases: 71.4%, controls: 75.0%).

**Table 1.**
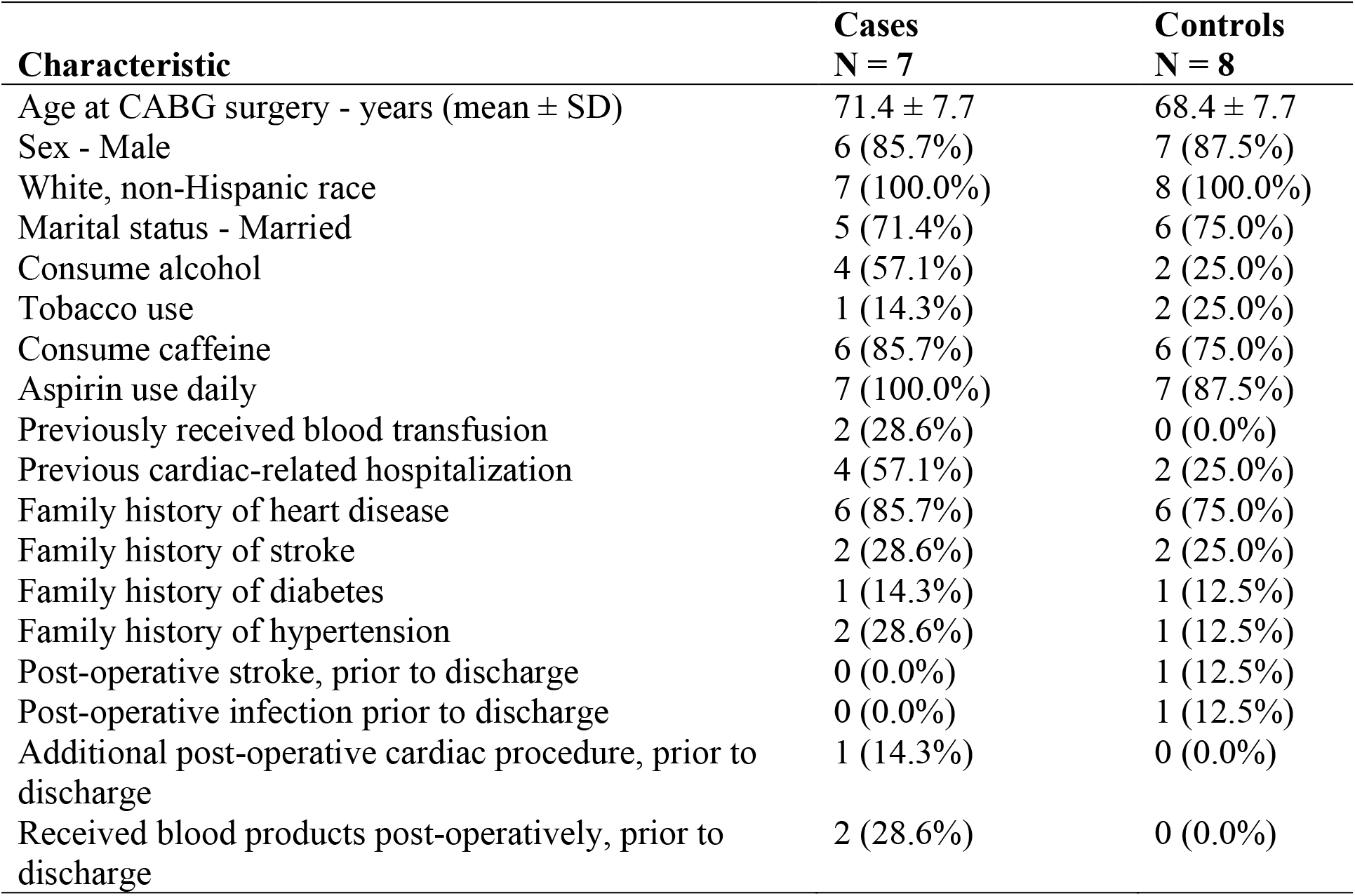
Baseline characteristics of the study population.

| **Characteristic** | **Cases**<br>**N = 7** | **Controls**<br>**N = 8** |
| --- | --- | --- |
| Age at CABG surgery - years (mean ± SD) | 71.4 ± 7.7 | 68.4 ± 7.7 |
| Sex - Male | 6 (85.7%) | 7 (87.5%) |
| White, non-Hispanic race | 7 (100.0%) | 8 (100.0%) |
| Marital status - Married | 5 (71.4%) | 6 (75.0%) |
| Consume alcohol | 4 (57.1%) | 2 (25.0%) |
| Tobacco use | 1 (14.3%) | 2 (25.0%) |
| Consume caffeine | 6 (85.7%) | 6 (75.0%) |
| Aspirin use daily | 7 (100.0%) | 7 (87.5%) |
| Previously received blood transfusion | 2 (28.6%) | 0 (0.0%) |
| Previous cardiac-related hospitalization | 4 (57.1%) | 2 (25.0%) |
| Family history of heart disease | 6 (85.7%) | 6 (75.0%) |
| Family history of stroke | 2 (28.6%) | 2 (25.0%) |
| Family history of diabetes | 1 (14.3%) | 1 (12.5%) |
| Family history of hypertension | 2 (28.6%) | 1 (12.5%) |
| Post-operative stroke, prior to discharge | 0 (0.0%) | 1 (12.5%) |
| Post-operative infection prior to discharge | 0 (0.0%) | 1 (12.5%) |
| Additional post-operative cardiac procedure, prior to discharge | 1 (14.3%) | 0 (0.0%) |
| Received blood products post-operatively, prior to discharge | 2 (28.6%) | 0 (0.0%) |

Important clinical features included a high prevalence of alcohol consumption among cases (57.1%) compared to controls (25.0%). Tobacco use was observed in 14.3% of cases versus 25.0% of controls. All cases (100%) and 87.5% of controls had used aspirin daily. Further clinical history characteristics and other pertinent variables are detailed in **Table 1**.

### Statistical analysis

Both cases and controls had similar mean ages with overlapping standard deviations. The t-test revealed no significant (p ≥ 0.05) age difference between the groups.

A chi-square analysis was conducted to assess the association between POAF and two categorical factors: alcohol consumption and tobacco use. The results indicated that alcohol consumption was not significantly associated with POAF, as the *p*-value was 0.46. Similarly, no significant relationship was observed between tobacco use and POAF, with a *p*-value of 1.0. The results suggested that none of these factors were significantly associated with POAF in this cohort. However, due to the limited sample size, further investigation with larger patient groups is warranted to validate these findings.

### Identifying the miRNAs predictive of AFib

To identify the miRNAs that are predictive of AFib, we employed a correlation-based feature selection method. We eliminated the features that were highly correlated (R <0.80) and selected the top 10 ranked miRNAs out the 84 miRNAs. We divided the dataset into training and validation sets (at a 7:3 ratio) and evaluated the performance of four machine learning methods—Random Forest, K-Nearest Neighbor (KNN), XGBoost, and Support Vector Machine (SVM)—in classifying AFib and control samples using the 10 selected miRNAs obtained from correlation-based feature selection. These 10 miRNAs, including hsa-miR-19a-3p, hsa-miR-19b-3p, hsa-miR-184, hsa-miR-200a-3p, hsa-let-7a-5p, hsa-miR-124-3p, hsa-miR-423-5p, hsa-miR-96-5p, hsa-miR-100-5p, and hsa-miR-17-5p, were collectively termed as an AFib-miRNA signature. The Random Forest model demonstrated perfect performance on the training dataset with an accuracy, sensitivity, specificity, and AUC of 1.0. However, its performance slightly dropped on the test dataset, with an accuracy, sensitivity, specificity, and an AUC of 0.80, 0.87, 0.71, and 0.76, respectively. The KNN model showed a balanced performance across both datasets. On the training dataset, it achieved an accuracy, sensitivity, specificity, and an AUC of 0.75, 0.62, 1.0, and 0.84, respectively. On the test dataset, it achieved an accuracy of 0.80, sensitivity of 0.62, specificity of 1.0, and an AUC of 0.77. XGBoost yielded high performance on both datasets. It achieved an accuracy, sensitivity, specificity, and an AUC of 0.9, 0.90, 0.89, and 0.97, respectively, on the training dataset. On the test dataset, it maintained a good accuracy, sensitivity, specificity, and an AUC of 0.73, 0.75, 0.71, and 0.83, respectively. The SVM model had the lowest performance among the evaluated methods, and obtained an accuracy, sensitivity, specificity, and an AUC of 0.55, 0.62, 0.46, and 0.53, respectively on the training dataset. Its performance further declined on the test dataset, as shown in **Table 2**.

**Table 2.** Prediction performance of the machine learning methods.

| **Method** | **Acc-Tr** | **Acc-Test** | **Se-Tr** | **Se-Test** | **Sp-Tr** | **Sp-Test** | **AUC-Tr** | **AUC-Test** |
| --- | --- | --- | --- | --- | --- | --- | --- | --- |
| Random Forest | 1 | 0.8 | 1 | 0.87 | 1 | 0.71 | 1 | 0.76 |
| KNN | 0.75 | 0.8 | 0.62 | 0.62 | 0.89 | 1 | 0.84 | 0.77 |
| XGBoost | 0.9 | 0.73 | 0.90 | 0.75 | 0.89 | 0.71 | 0.97 | 0.83 |
| SVM | 0.55 | 0.46 | 0.62 | 0.37 | 0.46 | 0.57 | 0.53 | 0.60 |

\*KNN: K-Nearest neighbor
\*Acc: Accuracy
\*Tr: Training
\*Se: Sensitivity
\*Sp: Specificity
\*AUC: Area Under the ROC Curve

Overall, XGBoost exhibited the highest AUC on both training and test datasets, indicating its superior performance in distinguishing AFib from control samples. These results underscore the importance of model selection and validation in the development of robust classifiers for AFib detection using miRNAs. The prediction performance of these models were evaluated using ROC curves, as shown in **Fig. 2**.

**Figure 2.**
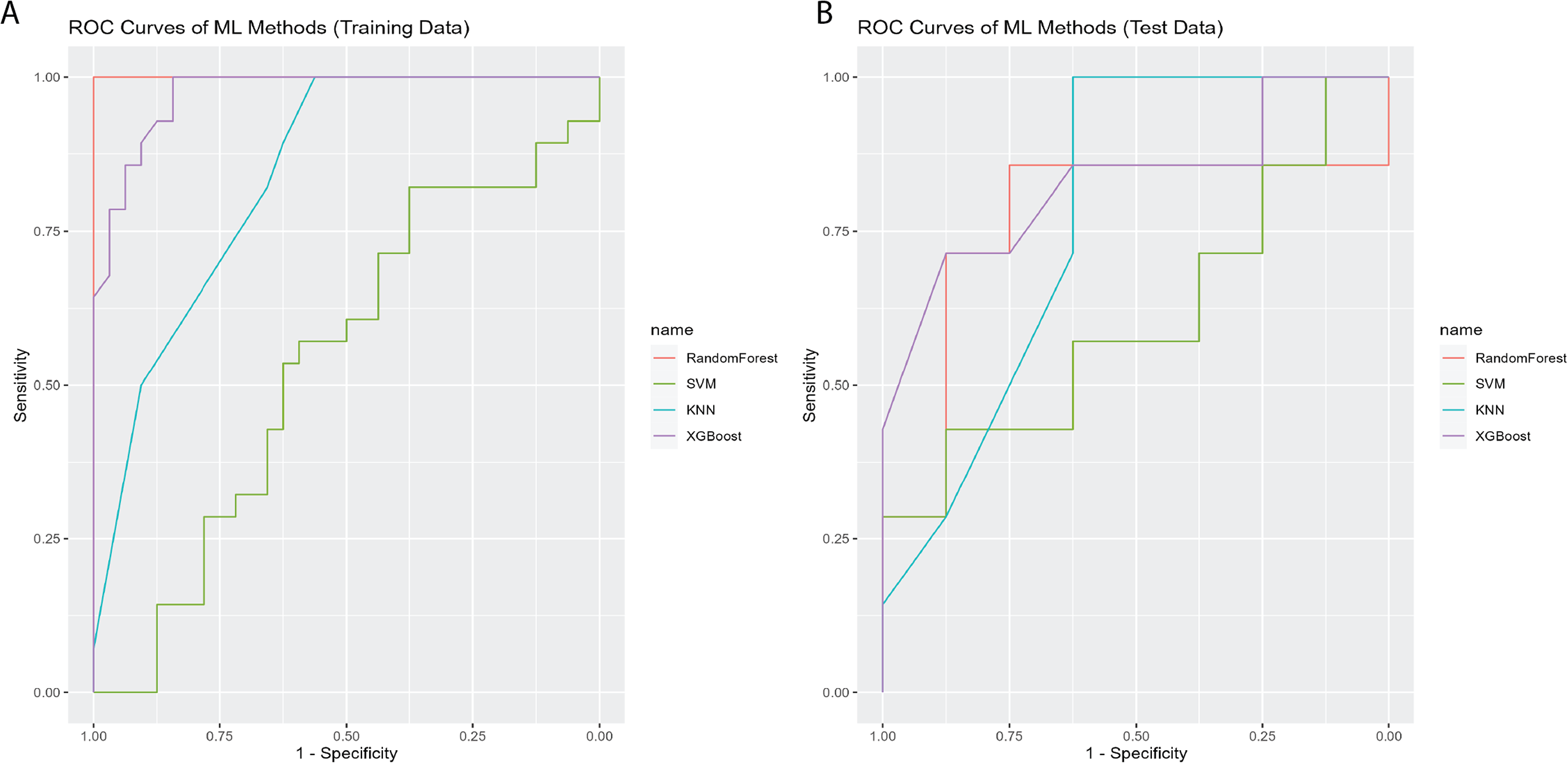
The comparison of prediction performance. (A) Evaluation of machine learning methods’ prediction ability using ROC curves to distinguish AFib patients from controls in the training dataset. (B) Evaluation using test datasets.

### Identification of differentially expressed miRNAs

Differential expression analysis (DEA) was employed to evaluate expression variations in the miRNA expression profiles of patients with atrial fibrillation (AFib) (n=7) and controls (n=8). Differentially expressed miRNAs were screened based on fold change and a FDR threshold of q<0.05. A total of 13 miRNAs were identified, with 12 being upregulated, including hsa-miR-96-5p, hsa-miR-184, hsa-miR-208a-3p, hsa-miR-17-5p, hsa-miR-499a-5p, hsa-miR-200a-3p, hsa-miR-203a, hsa-miR-210-3p, hsa-miR-9-5p, hsa-miR-204-5p, hsa-miR-146a-5p, and hsa-miR-133b. One miRNA, hsa-miR-574-3p, was downregulated, as shown in **Fig. 3A**. Four of these miRNAs—hsa-miR-96-5p, hsa-miR-184, hsa-miR-17-3p, and hsa-miR-200a-3p—overlap with the AFIB-miRNA signature (**Fig. 3B**).

**Figure 3.**
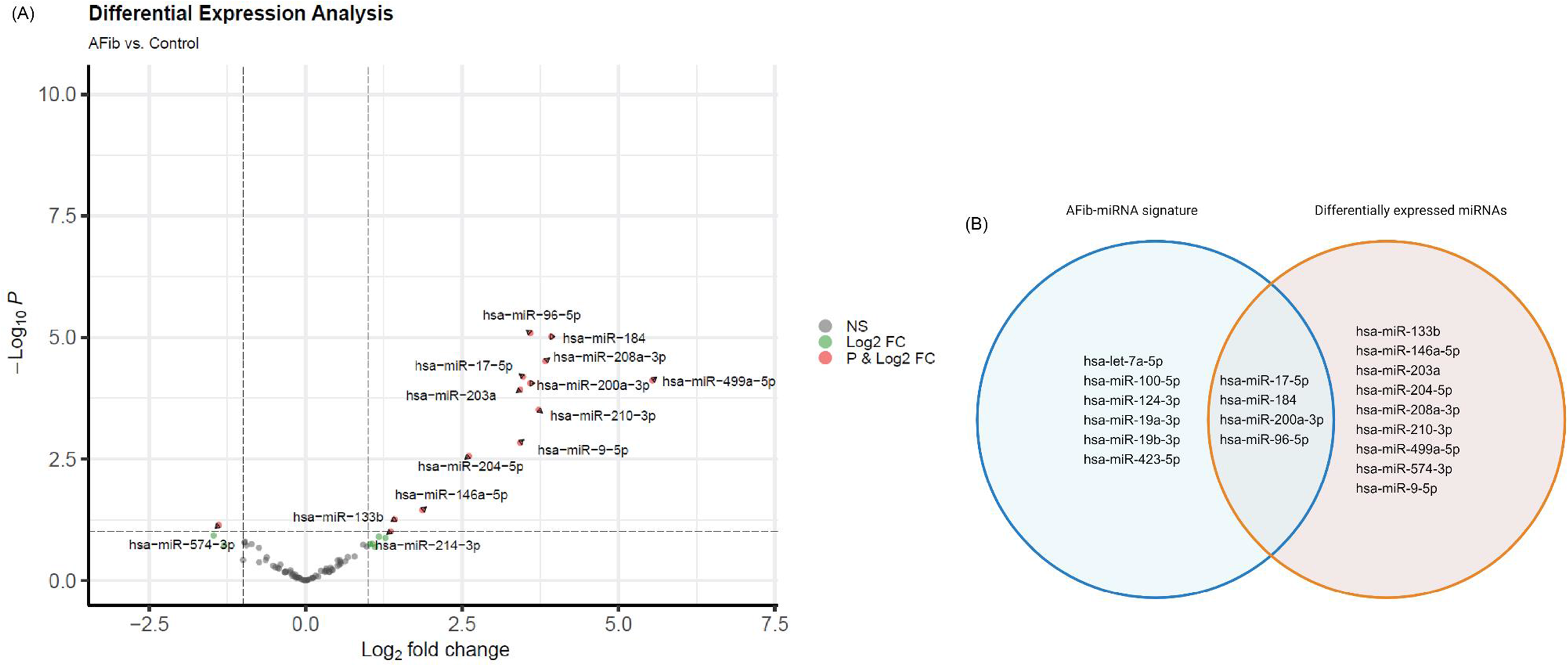
Differential expression analysis. (A) Volcano plot showing 12 upregulated and one downregulated miRNAs. (B) Venn diagram displaying the overlap between the 10 miRNAs in the AFib-miRNA signature identified by machine learning analysis and the 13 differentially expressed miRNAs.

### AFib-miRNAs and disease association

The AFib-miRNAs target various genes that can contribute to the development of disease pathways. These miRNAs are involved in numerous disease pathways, including cardiovascular diseases, cancers, and other non-cardiovascular diseases. The top 10 significant (*p*<0.001) non-cardiovascular diseases associated with these miRNAs include Vulvar Carcinoma, Multiple Sclerosis, Retinoblastoma, Kaposi’s Sarcoma, Medulloblastoma, Chronic Kidney Disease, Macular Degeneration, B-Cell Leukemia, Crohn’s Disease, and Myeloid Leukemia. MiRNAs such as hsa-let-7a-5p, hsa-miR-96-5p, hsa-miR-124-5p, and hsa-miR-17-5p showed enrichment in multiple diseases (**Fig. 4A**), suggesting their broader impact on disease pathology. The majority of the diseases associated with these miRNAs are related to cancer.

**Figure 4.**
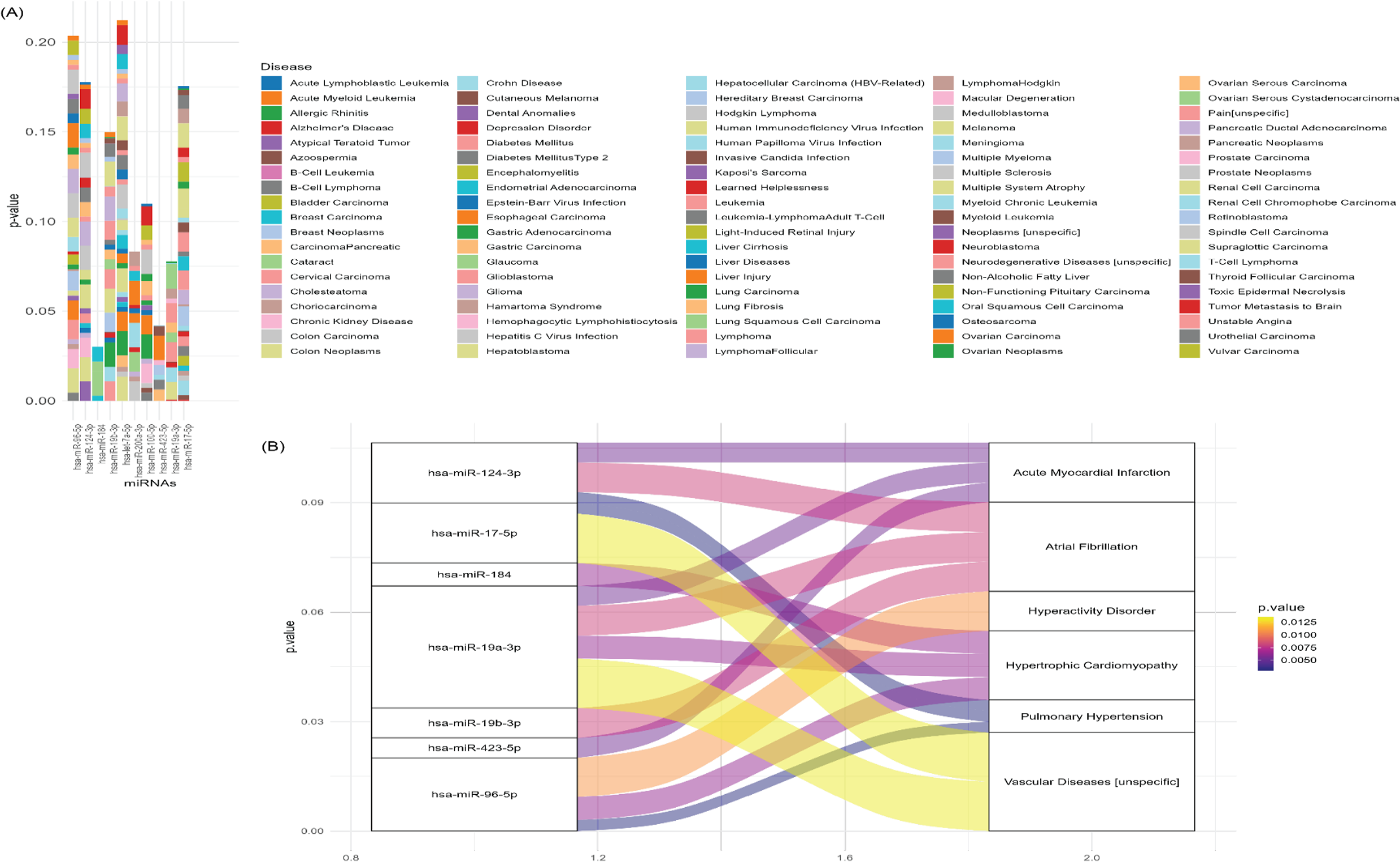
MiRNA-disease association. (A) Bar plot showing the association of the AFib-miRNA signature with various cancers and diseases. The X-axis represents AFib-miRNAs and the Y-axis shows p-values. (B) Alluvial plot illustrating the AFib-miRNA signature association specific to cardiovascular diseases.

Furthermore, we identified associations between seven miRNAs in the AFib-miRNA signature and cardiovascular diseases. The miRNAs hsa-miR-124-3p, hsa-miR-17-5p, hsa-miR-184, hsa-miR-19a-3p, hsa-miR-19b-3p, hsa-miR-423-5p, and hsa-miR-96-5p showed statistically significant (*p*<0.005) associations with AFib, acute myocardial infarction, hyperactivity disorder, hypertrophic cardiomyopathy, pulmonary hypertension, and vascular disease, as shown in **Fig. 4B**.

### Biological relevance of the AFib-miRNA signature

We investigated the biological relevance of the miRNAs using KEGG, GO, Reactome pathway analysis. The AFib-miRNA signature play crucial roles in various biological functions, impacting multiple diseases, including cardiovascular diseases. In KEGG pathway analysis, the AFib-miRNA signature was significantly enriched in pathways such as proteoglycans in cancer, p53 signaling pathway, hepatitis B, EGFR tyrosine kinase inhibitor resistance, and AGE-RAGE signaling pathway in diabetic complications, as detailed in **Supplementary Table 1**. The enrichment analysis of the AFib-miRNA signature is illustrated in **Supplementary Fig. 1A**. Focusing on AFib-miRNA signature enriched in cardiovascular-related pathways, we identified specific KEGG pathways involved in cardiovascular diseases, including hypertrophic cardiomyopathy, MAPK signaling, PI3K-Akt signaling, FoxO signaling, and TGF-beta signaling (**Table 3** and **Supplementary Fig. 1B**).

**Table 3.** Involvement of AFib-miRNA signature in cardiovascular-associated pathways.

| **Term Name** | **miRNAs** | **P-value** | **FDR** |
| --- | --- | --- | --- |
| Hypertrophic cardiomyopathy | hsa-miR-17-5p,hsa-miR-19a-3p | 9.32E-14 | 1.55E-13 |
| Dilated cardiomyopathy | hsa-miR-17-5p,hsa-miR-19a-3p | 1.87E-14 | 3.28E-14 |
| MAPK signaling pathway | hsa-let-7a-5p,hsa-miR-17-5p,hsa-miR-19a-3p | 2.89E-18 | 7.04E-18 |
| PI3K-Akt signaling pathway | hsa-let-7a-5p,hsa-miR-17-5p,hsa-miR-124-3p,hsa-miR-200a-3p | 2.00E-23 | 1.05E-22 |
| FoxO signaling pathway | hsa-let-7a-5p,hsa-miR-17-5p,hsa-miR-19a-3p,hsa-miR-200a-3p | 3.34E-27 | 3.34E-26 |
| TGF-beta signaling pathway | hsa-miR-17-5p,hsa-miR-19a-3p | 9.43E-27 | 7.17E-26 |
| Fluid shear stress and atherosclerosis | hsa-miR-17-5p,hsa-miR-19a-3p | 5.56E-20 | 1.69E-19 |
| Adrenergic signaling in cardiomyocytes | hsa-miR-19a-3p | 0.0127445 | 0.013048 |
| Vascular smooth muscle contraction | hsa-miR-124-3p | 0.013172 | 0.013406 |
| Viral myocarditis | hsa-miR-423-5p | 0.001448 | 0.001754 |
| cGMP-PKG signaling pathway | hsa-miR-184 | 0.0022891 | 0.002734 |
| HIF-1 signaling pathway | hsa-let-7a-5p,hsa-miR-96-5p,hsa-miR-124-3p,hsa-miR-184 | 1.26E-10 | 3.34E-10 |

GO annotation analysis revealed a strong association between the miRNAs and biological processes, such as the positive regulation of transcription by RNA polymerase II, the regulation of gene expression (both positive and negative), cytokine-mediated signaling, regulation of the apoptotic process, and heart development. Significant GO terms (Benjamini-Hochberg FDR *q* < 0.001) are shown in **Fig. 5A**, with detailed miRNA enrichment in biological processes and *p*-values listed in **Supplementary Table S2**. GO molecular functions analysis highlighted that the AFib miRNA signature is highly enriched in transcription binding, DNA-binding transcription factor activity, RNA polymerase II-specific activity, protein binding, RNA polymerase II-cis-regulatory region sequence-specific DNA binding, and protease binding (**Fig. 5B**). Detailed enrichment in molecular functions, target genes, and p-values are provided in **Supplementary Table S3**. In terms of cellular components, the AFib miRNA signature is enriched in the nucleoplasm, nucleus, nuclear chromatin, membrane raft, and cytoplasm (**Fig. 5C**), with details in **Supplementary Table S4**.

**Figure 5.**
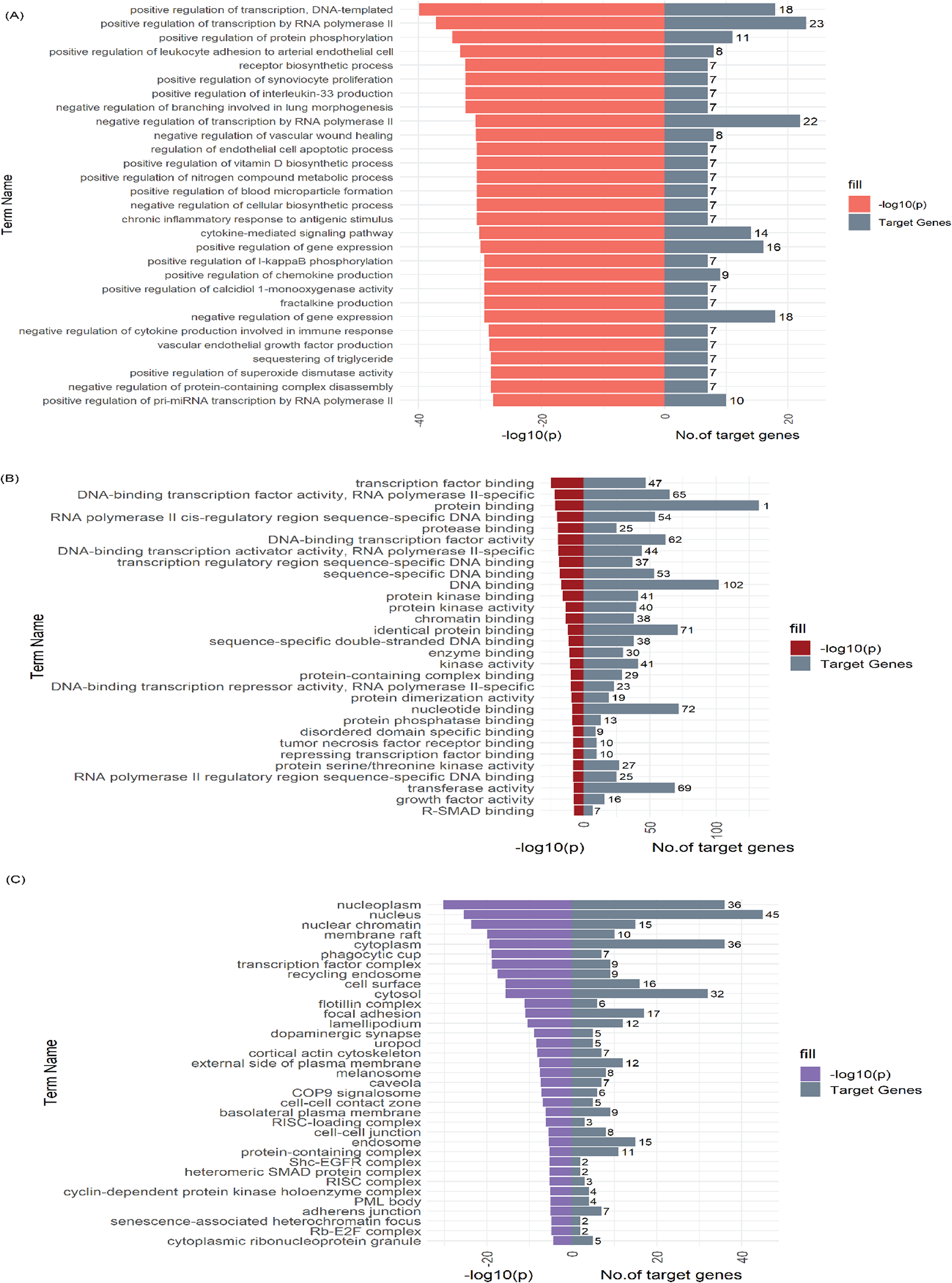
AFib-miRNA signature enrichment in gene ontology annotations. (A) Bar plots showing the enrichment of the AFib-miRNA signature in biological processes. (B) Enrichment in molecular functions. (C) Enrichment in cellular components. The X-axis represents –log10 (p-value) and the Y-axis shows the number of target genes and Gene Ontology term names.

We further identified GO categories specific to cardiovascular diseases. The AFib-miRNA signature are significantly enriched in biological processes including heart development, positive regulation of smooth muscle cell proliferation, striated muscle cell differentiation, cortical actin cytoskeleton organization, and positive regulation of leukocyte adhesion to arterial endothelial cells (**Fig. 6A**). For molecular functions, the AFib-miRNA signature is enriched in chromatin binding, DNA binding transcription factor activity, enzyme binding, identical protein binding, and protein kinase activity (**Fig. 6B**). In cellular components, enrichments include caveola, cell-cell junction, COP9 signalosome, cytoplasm, and dopaminergic synapse (**Fig. 6C**). Detailed information on AFib-miRNA signature enrichment in cardiovascular-related GO categories, target genes, and p-values is available in **Supplementary Table S5**.

**Figure 6.**
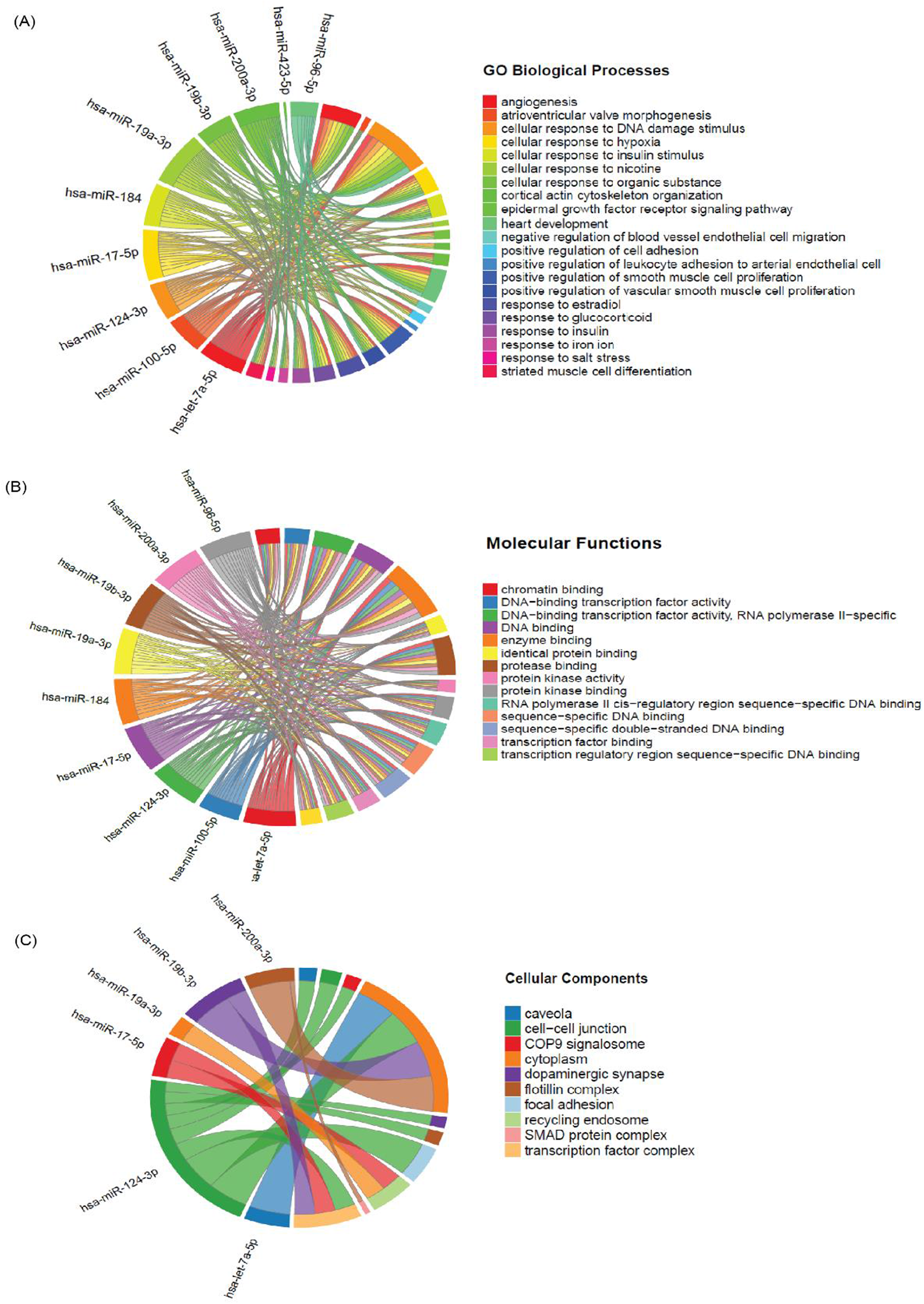
AFib-miRNA signature in cardiovascular-associated Gene Ontology annotations. **(A) Chord diagram displaying the involvement of the AFib-miRNA signature in biological processes. (B) Involvement in molecular functions. (C) Involvement in cellular components.**

Additionally, the AFib miRNA signature enriched in Reactome pathways including Interleukin-4 and Interleukin-13 signaling, TNFR1-mediated ceramide production, TNFR1-induced proapoptotic signaling, TNFR1-induced NFkappaB signaling pathway, and Interleukin-10 signaling, as shown in **Supplementary Table S6**.

### AFib-miRNA-gene interaction prediction

We identified key gene targets associated with the AFib-miRNA signature to uncover the genetic network affected by these miRNAs and provide insights into the biological alterations linked to cardiovascular diseases. Using miRWalk, miRNet, miRDB, and miRTarBase, we identified 9,763 gene targets for the AFib miRNA signature. We focused on gene targets shared by five or more of the AFib-miRNA signature miRNAs (**Fig. 7A**). This approach led us to identify 50 genes. Next, we used miRNet (2.0) [22] to construct a miRNA-gene interaction network with 30 gene targets using the shortest path algorithm (**Fig. 7B**). We examined the expression of these 30 genes in human dilated and hypertrophic cardiomyopathy. This analysis revealed that these genes are enriched in fibroblasts, cardiomyocyte-I, II, and III, macrophages, and adipocytes (**Fig. 7C**). Furthermore, we built a miRNA-small molecule interaction network to explore the targets for the AFib miRNA signature. The AFib-miRNA signature targets various small molecules, as shown in **Fig. 7D**.

**Figure 7.**
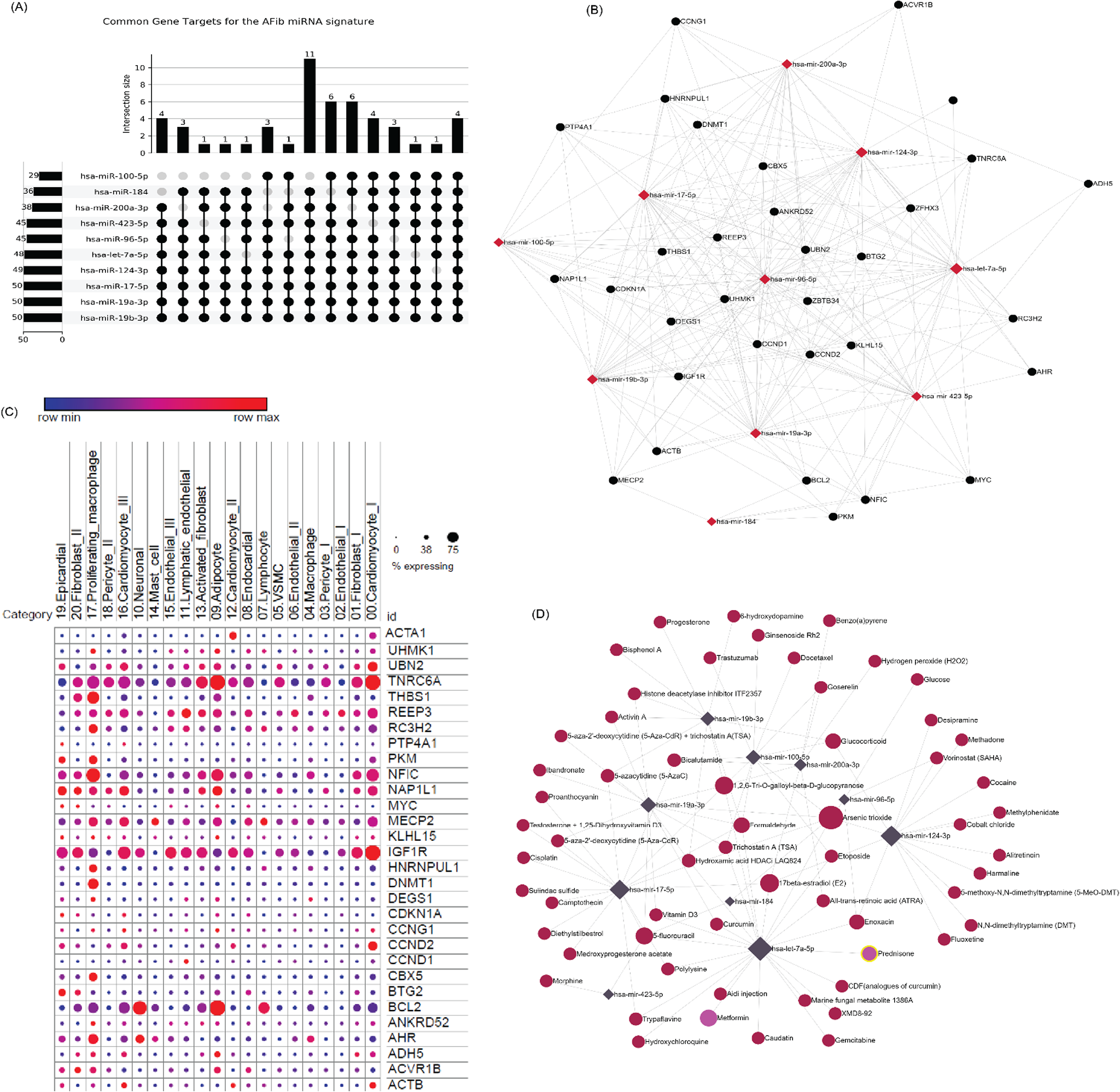
MiRNA-gene target interactions. (A) Upset plot depicting shared gene targets within the AFib-miRNA signature. (B) Network demonstrating connections between AFib-miRNAs (in red rhombus) and target genes (in black circles). (C) Comparative expression levels of AFib-miRNA signature targeted genes in hypertrophic cardiomyopathy. Plot generated from the Broad Institute of MIT and Harvard’s Single Cell Portal. (D) Network plot displaying specific predicted small molecule drugs targeting the miRNA signature, with miRNAs represented in navy blue rhombus and small molecules in burgundy circles.

## Discussion

Early detection of AFib holds significant promise as it can inform management decisions that alter the natural progression and complication profile of this disease. Traditional parameters, including demographic characteristics such as age, gender, alcohol consumption, tobacco use, and previous medical history, may not reliably predict AF, especially in studies with small sample sizes. However, molecular information may offer valuable insights.

In this study, we investigated the presence of circulating miRNAs in patients who had undergone CABG by employing microarray and bioinformatics analyses. These techniques allowed us to identify potential biomarkers for predicting AFib post-CABG. Leveraging machine learning techniques, we selected the top 10 ranked miRNAs out of 84 and evaluated their AFib prediction performance using Random Forest, KNN, XGBoost, and SVM models. The Random Forest and XGBoost models demonstrated superior prediction performance compared to the other methods, with test sensitivities of 0.76 and 0.83, respectively. To further explore the miRNAs differentially expressed between cases and controls, we performed differential expression analysis. We found that four of these upregulated miRNAs—hsa-miR-96-5p, hsa-miR-184, hsa-miR-17-3p, and hsa-miR-200-3p—overlapped with the AF-miRNA signature. While the 13 differentially expressed miRNAs showed significant differences between cases and controls, nine of the 13 miRNAs were not accurately predictive of AF. Thus, we focused exclusively on the AF-miRNA signature for further analysis. It is important to note that our preliminary analysis, previously published, identified a different set of miRNAs and primarily involved statistical analysis[13]. The findings from our current analysis, focusing on miRNA identification, are detailed in this study.

The association between the AFib-miRNA signature and cardiovascular diseases revealed that miRNAs such as hsa-miR-124-3p, hsa-miR-17-5p, hsa-miR-184, hsa-miR-19a-3p, hsa-miR-19b-3p, hsa-miR-423-5p, and hsa-miR-96-5p were significantly associated with various cardiovascular diseases, including acute myocardial infarction, hyperactivity disorder, hypertrophic cardiomyopathy, pulmonary hypertension, and vascular disease. The literature supports these findings. For instance, miR-124-3p promotes cardiac fibroblast activation, while miR-17-5p is associated with acute myocardial ischemia injury and cardiac hypertrophy and serves as a novel biomarker for diagnosing acute myocardial infarction [27], while miR-17-5p is associated with acute myocardial ischemia injury and cardiac hypertrophy and serves as a novel biomarker for diagnosing acute myocardial infarction [28–30]. Circulating miR-184 is a potential predictive biomarker for cardiac damage and acts as a biomarker for AFib in patients with valvular heart disease [12]. Lower expression levels of miR-19a-3p/19b-3p are found in the plasma of heart failure patients, with miR-19b-3p identified as a strong prognostic biomarker for acute heart failure [31, 32]. Increased expression levels of miR-423-5p are linked to heart failure diagnosis [33], and miR-96-5p functions as a potential diagnostic biomarker for acute myocardial infarction [34].

Further biological relevance of the AFib-miRNAs revealed that these miRNAs target specific KEGG pathways associated with cardiovascular diseases, such as hypertrophic cardiomyopathy (HCM), MAPK signaling pathway, PI3K-Akt signaling pathway, FoxO signaling pathway, and TGF-beta signaling pathway. AFib is a common sequela of HCM, with evidence showing approximately a 20% lifetime risk for the development of AFib in HCM [35, 36]. The HCM pathway maps out the genetic and metabolic interactions involved in the disease. Hundreds of gene mutations in the sarcomere proteins are linked to HCM, increasing the Ca2+ sensitivity of cardiac myofilaments. This heightened sensitivity likely raises ATP utilization, potentially causing an energy imbalance in the heart under stress. Specific miRNAs such as hsa-miR-17-5p and hsa-miR-19a-3p influence key signaling molecules and pathways in HCM, including those involved in cardiac hypertrophy and fibrosis [37–39]. For example, miR-19a-3p and 19b-3p target components of the TGF-β signaling pathway, crucial in the fibrotic response seen in HCM [39]. KEGG pathway analysis showed that the AFib-miRNAs, including hsa-let-7a-5p, hsa-miR-17-5p, hsa-miR-19a-3p, hsa-miR-124-3p, and hsa-miR-200-3p, were significantly (P<0.005) involved in the MAPK signaling pathway, PI3K-Akt signaling pathway, and FoxO signaling pathway. These pathways are important in atrial fibrosis in patients with chronic AFib [40–43].

Gene Ontology (GO) annotations provide key insights into the biological processes, cellular components, and molecular functions associated with AFib. AFib-miRNAs play a significant role in AFib by regulating the expression of genes involved in crucial pathways such as heart development and positive regulation of smooth muscle cell proliferation, contributing to the pathogenesis of AFib [44–47]. GO annotation analysis also provides insights into the enrichment of AFib-miRNAs in molecular functions, including transcription binding, DNA-binding transcription factor activity, and RNA polymerase II-specific; cellular components such as caveolae, cell-cell junction, COP9 signalosome, and cytoplasm. This comprehensive analysis highlights the importance of the identified AFib-miRNAs in AFib development and cardiovascular disease progression.

The identified AFib-miRNA signature has the potential to be predictive of POAF in patients undergoing CABG as shown by its significant enrichment in several important pathways that contribute to AFib and other cardiovascular diseases. However, this study has some limitations. The small sample size limits the generalizability and robustness of the findings, necessitating further validation with larger cohorts. Additionally, while the miRNA signatures identified show promise, their predictive power needs to be confirmed through independent validation studies. The study also does not account for potential confounding factors such as medication use, which might influence miRNA expression. Future research should aim to address these limitations by incorporating larger, diverse populations and considering additional variables that may affect miRNA expression.

## Data availability

Data in a de-identified format will be made available by request to the corresponding author.

## Data Availability

All data are provided in the manuscript and supplemental information.

## Acknowledgement

This work was supported in part by the Marshfield Clinic Research Institute, Marshfield, WI. The funders had no role in the study design, data collection and analysis, decision to publish, or preparation of the manuscript.

## Author contributions

S.Y.S and P.S. supervised and carried out the detail study. S.Y.S, T.C, D.S, N.S, S.K.S, J.P, G.I, J.B and P.S participated in data analysis, manuscript preparation and discussed the results. All authors have read and approved the final manuscript.

## Supplemental Information

Supplementary file contains Figure and Tables.

Supplementary Figure and Table legends

**Supplementary Figure S1**. Enrichment of AFib-miRNAs in KEGG Pathways. (A) MiRNA enrichment in various cancer and disease pathways. (B) MiRNA involvement in cardiovascular-specific pathways.

**Supplementary Table S1**. Enrichment of AFib-miRNA signature in KEGG pathways.

**Supplementary Table S2**. Enrichment of AFib-miRNA signature in gene ontology biological processes.

**Supplementary Table S3**. Enrichment of AFib-miRNA signature in molecular functions.

**Supplementary Table S4**. Enrichment of AFib-miRNA signature in cellular components.

**Supplementary Table S5**. Enrichment of AFib-miRNA signature in Gene Ontology categories specific to cardiovascular diseases.

**Supplementary Table S6**. Enrichment of the AFib-miRNA signature in Reactome pathways.

## References

1. Everett, T.H.t. and J.E. Olgin, Atrial fibrosis and the mechanisms of atrial fibrillation. Heart Rhythm, 2007. 4(3 Suppl): p. S24–7.

2. Auer, J., et al., Risk factors of postoperative atrial fibrillation after cardiac surgery. J Card Surg, 2005. 20(5): p. 425–31.

3. Mathew, J.P., et al., A multicenter risk index for atrial fibrillation after cardiac surgery. Jama, 2004. 291(14): p. 1720–9.

4. Fuster, V., et al., 2011 ACCF/AHA/HRS focused updates incorporated into the ACC/AHA/ESC 2006 Guidelines for the management of patients with atrial fibrillation: a report of the American College of Cardiology Foundation/American Heart Association Task Force on Practice Guidelines developed in partnership with the European Society of Cardiology and in collaboration with the European Heart Rhythm Association and the Heart Rhythm Society. J Am Coll Cardiol, 2011. 57(11): p. e101–98.

5. Sun, Y., et al., Role of preoperative atorvastatin administration in protection against postoperative atrial fibrillation following conventional coronary artery bypass grafting. Int Heart J, 2011. 52(1): p. 7–11.

6. Attaran, S., et al., Atrial fibrillation postcardiac surgery: a common but a morbid complication. Interact Cardiovasc Thorac Surg, 2011. 12(5): p. 772–7.

7. Baker, W.L. and C.M. White, Post-cardiothoracic surgery atrial fibrillation: a review of preventive strategies. Ann Pharmacother, 2007. 41(4): p. 587–98.

8. Elahi, M.M., S. Flatman, and B.M. Matata, Tracing the origins of postoperative atrial fibrillation: the concept of oxidative stress-mediated myocardial injury phenomenon. Eur J Cardiovasc Prev Rehabil, 2008. 15(6): p. 735–41.

9. Chung, M.K., et al., C-reactive protein elevation in patients with atrial arrhythmias: inflammatory mechanisms and persistence of atrial fibrillation. Circulation, 2001. 104(24): p. 2886–91.

10. Baba, A. and M. Fu, Autoantibodies in atrial fibrillation: actor, biomaker or bystander? Autoimmunity, 2008. 41(6): p. 470–2.

11. Andrew, P. and A.S. Montenero, Is there a link between atrial fibrillation and certain bacterial infections? J Cardiovasc Med (Hagerstown), 2007. 8(12): p. 990–6.

12. Sharma Param, P., et al., IDENTIFICATION OF A MICRORNA SIGNATURE FOR PREDICTING ATRIAL FIBRILLATION IN PATIENTS WITH VALVULAR HEART DISEASE. Journal of the American College of Cardiology, 2024. 83(13_Supplement): p. 2185–2185.

13. Sathipati Srinivasulu, Y., et al., CIRCULATING MICRORNA SIGNATURE PREDICTS ATRIAL FIBRILLATION IN PATIENTS FOLLOWING CORONARY ARTERY BYPASS GRAFTING. Journal of the American College of Cardiology, 2024. 83(13_Supplement): p. 33–33.

14. Gilad, S., et al., Serum microRNAs are promising novel biomarkers. PLoS One, 2008. 3(9): p. e3148.

15. Barth, A.S., et al., Reprogramming of the human atrial transcriptome in permanent atrial fibrillation: expression of a ventricular-like genomic signature. Circ Res, 2005. 96(9): p. 1022–9.

16. Shen, N.N., et al., Identification of microRNA biomarkers in atrial fibrillation: A protocol for systematic review and bioinformatics analysis. Medicine (Baltimore), 2019. 98(30): p. e16538.

17. Koniari, I., et al., Biomarkers in the clinical management of patients with atrial fibrillation and heart failure. J Geriatr Cardiol, 2021. 18(11): p. 908–951.

18. Galenko, O., et al., The role of microRNAs in the development, regulation, and treatment of atrial fibrillation. Journal of Interventional Cardiac Electrophysiology, 2019. 55(3): p. 297–305.

19. Kiliszek, M., et al., Serum microRNA in patients undergoing atrial fibrillation ablation. Scientific Reports, 2020. 10(1): p. 4424.

20. Cao, Y. and L. Cui, Identifying the key microRNAs implicated in atrial fibrillation. Anatol J Cardiol, 2021. 25(6): p. 429–436.

21. Cui, C., et al., HMDD v4.0: a database for experimentally supported human microRNA-disease associations. Nucleic Acids Research, 2024. 52(D1): p. D1327–D1332.

22. Chang, L. and J. Xia, MicroRNA Regulatory Network Analysis Using miRNet 2.0. Methods Mol Biol, 2023. 2594: p. 185–204.

23. Tastsoglou, S., et al., DIANA-miRPath v4.0: expanding target-based miRNA functional analysis in cell-type and tissue contexts. Nucleic Acids Research, 2023. 51(W1): p. W154–W159.

24. Sticht, C., et al., miRWalk: An online resource for prediction of microRNA binding sites. PLoS One, 2018. 13(10): p. e0206239.

25. Chen, Y. and X. Wang, miRDB: an online database for prediction of functional microRNA targets. Nucleic Acids Res, 2020. 48(D1): p. D127–d131.

26. Huang, H.Y., et al., miRTarBase update 2022: an informative resource for experimentally validated miRNA-target interactions. Nucleic Acids Res, 2022. 50(D1): p. D222–d230.

27. Zhu, P., et al., MicroRNAs sequencing of plasma exosomes derived from patients with atrial fibrillation: miR-124-3p promotes cardiac fibroblast activation and proliferation by regulating AXIN1. J Physiol Biochem, 2022. 78(1): p. 85–98.

28. Zhao, L., et al., MiR-17-5p-mediated endoplasmic reticulum stress promotes acute myocardial ischemia injury through targeting Tsg101. Cell Stress Chaperones, 2021. 26(1): p. 77–90.

29. Xu, X., et al., MicroRNA-17-5p Promotes Cardiac Hypertrophy by Targeting Mfn2 to Inhibit Autophagy. Cardiovasc Toxicol, 2021. 21(9): p. 759–771.

30. Xue, S., et al., Circulating MiR-17-5p, MiR-126-5p and MiR-145-3p Are Novel Biomarkers for Diagnosis of Acute Myocardial Infarction. Front Physiol, 2019. 10: p. 123.

31. Zou, M., et al., Autophagy inhibition of hsa-miR-19a-3p/19b-3p by targeting TGF-β R II during TGF-β1-induced fibrogenesis in human cardiac fibroblasts. Scientific Reports, 2016. 6(1): p. 24747.

32. Su, Y., et al., Circulating miR-19b-3p as a Novel Prognostic Biomarker for Acute Heart Failure. Journal of the American Heart Association, 2021. 10(20): p. e022304.

33. Tijsen, A.J., et al., MiR423-5p As a Circulating Biomarker for Heart Failure. Circulation Research, 2010. 106(6): p. 1035–1039.

34. Ding, H., W. Chen, and X. Chen, Serum miR-96-5p is a novel and non-invasive marker of acute myocardial infarction associated with coronary artery disease. Bioengineered, 2022. 13(2): p. 3930–3943.

35. Siontis, K.C., et al., Atrial Fibrillation in Hypertrophic Cardiomyopathy: Prevalence, Clinical Correlations, and Mortality in a Large High-Risk Population. Journal of the American Heart Association. 3(3): p. e001002.

36. Olivotto, I., et al., Impact of atrial fibrillation on the clinical course of hypertrophic cardiomyopathy. Circulation, 2001. 104(21): p. 2517–2524.

37. Sun, Y., et al., Susceptibility modules and genes in hypertrophic cardiomyopathy by WGCNA and ceRNA network analysis. Frontiers in Cell and Developmental Biology, 2022. 9: p. 822465.

38. Shi, H., et al., Systematic identification and analysis of dysregulated mi RNA and transcription factor feed-forward loops in hypertrophic cardiomyopathy. Journal of Cellular and Molecular Medicine, 2019. 23(1): p. 306–316.

39. Zou, M., et al., Autophagy inhibition of hsa-miR-19a-3p/19b-3p by targeting TGF-β R II during TGF-β1-induced fibrogenesis in human cardiac fibroblasts. Scientific reports, 2016. 6(1): p. 24747.

40. Zhang, D., et al., Role of the MAPKs/TGF-β1/TRAF6 signaling pathway in postoperative atrial fibrillation. PLoS One, 2017. 12(3): p. e0173759.

41. Zhang, D., et al., Role of the MAPKs/TGF-β1/TRAF6 signaling pathway in atrial fibrosis of patients with chronic atrial fibrillation and rheumatic mitral valve disease. Cardiology, 2014. 129(4): p. 216–223.

42. McMullen, J.R., et al., Ibrutinib increases the risk of atrial fibrillation, potentially through inhibition of cardiac PI3K-Akt signaling. Blood, The Journal of the American Society of Hematology, 2014. 124(25): p. 3829–3830.

43. Yu, W., C. Chen, and J. Cheng, The role and molecular mechanism of FoxO1 in mediating cardiac hypertrophy. ESC Heart Failure, 2020. 7(6): p. 3497–3504.

44. Wang, D., et al., Long non-coding RNA MALAT1 sponges miR-124-3p. 1/KLF5 to promote pulmonary vascular remodeling and cell cycle progression of pulmonary artery hypertension. International journal of molecular medicine, 2019. 44(3): p. 871–884.

45. Sharma, V., et al., Integrative experimental validation of concomitant miRNAs and transcription factors with differentially expressed genes in acute myocardial infarction. European Journal of Pharmacology, 2024. 971: p. 176540.

46. Vatan, M.B., et al., Altered plasma microRNA expression in patients with mitral chordae tendineae rupture. J. Heart Valve Dis, 2016. 25: p. 580–588.

47. Su, Y., et al., Circulating miR-19b-3p as a Novel Prognostic Biomarker for Acute Heart Failure. Journal of the American Heart Association, 2021. 10(20): p. e022304.

